# A Systematic Review and Meta-analysis for Association of Celiac Disease and Thyroid Disorders

**DOI:** 10.1101/2024.01.26.24301845

**Authors:** Zahra Norouzi, Fatemeh Hasani, Sima Besharat, Hesamaddin Shirzad-Aski, Somayeh Ghorbani, Masoud Mohammadi, Anahita Yadegari, Ali Kalhori

## Abstract

It is probable that people who have celiac disease (CD) are more likely to have thyroid disorders. A comprehensive systematic review and meta-analysis were conducted to assess the link between thyroid disorders and CD. Articles were selected from PubMed, Web of Science, Scopus, Ovid, Embase, Cochrane, ProQuest, and Wiley from February 2022 and earlier. A meta-analysis was conducted to evaluate the outcomes, using odds ratios (ORs) and corresponding 95% confidence intervals (95% CIs). The meta-analysis comprised 31 articles with 3310256 participants including 101253 individuals with thyroid disorders. Overall, the frequency of thyroid disease was notably higher in patients with CD compared to the control groups (OR: 3.06, 95% CI: 2.51 – 3.72, P<0.001). The findings of our meta-analysis support the notion that patients with CD are more likely to have autoimmune thyroid disease (ATD) and other thyroid disorders than the control group, thus indicating that regular screening for thyroid disease is necessary for CD patients. Further cohort research is required to investigate the relationship between thyroid disorders and CD.

## Introduction

Celiac disease (CD) is an inflammatory autoimmune disorder of the upper small intestine. The disease triggered by gluten-containing food and environmental factors in genetically predisposed patients with the HLA-DQ2 and HLA-DQ8 (1). It affects approximately 1% of the global population (2).

CD patients are considered to have more autoimmune diseases than general population (3). One of the most frequent autoimmune conditions related to CD is autoimmune thyroid disease (ATD). ATD is a group of inflammatory disorders that affect the thyroid gland. The two most common forms of ATD are Graves’ disease and Hashimoto’s thyroiditis (4). These conditions are characterized by the presence of antibodies such as anti-thyroid peroxidase (TPO), anti-thyroglobulin (TG), and anti-thyroid-stimulating hormone (TSH) receptor antibodies (5). The risk for the development of these diseases is higher among women compared to men and it also increases with age (6, 7).

Graves’ disease is the most frequent cause of hyperthyroidism. This disorder is characterized by an autoimmune reaction against thyroid antigens, which leads to secretion of anti-TSH receptor autoantibodies by B cells. These autoantibodies bind to the TSH receptors and chronically stimulate them, leading to chronical secretion of thyroid hormones and thyrotoxicosis (6). On the other hand, Hashimoto’s thyroiditis can cause hypothyroidism and is clinically characterized by anti-TPO and anti-TG antibodies. In Hashimoto’s thyroiditis, increased infiltration of B and T cells into the thyroid gland causes destruction of follicular cells responsible for the production of thyroid hormones, resulting in hypothyroidism (7).

Although the precise reason for the co-occurrence of celiac disease and thyroid disorders is not yet fully understood, several theories have been suggested. These include genetic predisposition, similar cytokine pathways involved in both disease progression, and impaired absorption of key nutrients due to changes in gut permeability. Such factors may contribute to the higher incidence of these conditions occurring together (8).

There are several studies that showed this co-occurrence connection. In a cross-sectional study conducted on 288 CD patients, Baharvand et al. reported that thyroid disease was four times more prevalent in CD patients than in controls (9). Also, Norström, in a study of 335 children with CD, claimed that the risk of thyroid dysfunction associated with thyroid autoimmunity is higher for the CD patients than the healthy control (10). Obaid et al. also investigated 86 CD patients in Iraq from 2018 until 2020. The findings of the study showed a strong association between CD and ATDs (4). In addition, Bibbò et al. found a significant association between Hashimoto thyroiditis and CD, as the most prevalent autoimmune disease in CD patients than controls. (24.3% vs. 10%) (11).

In 2016, Sun et al. conducted a systematic review and meta-analysis comprising of 13 articles with a total of 15,629 CD cases and 79,342 controls. They discovered that the prevalence of thyroid disease was significantly higher in individuals with CD than in the control groups. The odds ratio (OR) was calculated to be 3.08 with a 95% confidence interval (CI) ranging from 2.67-3.56, indicating statistical significance with a P-value of less than 0.001. Interestingly, the analysis revealed that there was no significant difference in the OR between the groups that underwent gluten treatment and those who did not, with an OR of 1.08 and a 95% CI range of 0.61-1.92 (P=0.786). These findings suggest that individuals with CD are more likely to develop thyroid disorders and therefore should undergo regular screening for thyroid disease. (12).

However, there is still a need for a comprehensive and systematic review of the available literature to evaluate this association accurately. Therefore, and to clarify more, we did a new systematic review aimed at evaluating the incidence of thyroid disease in patients with CD after 5 years of the previous meta-analysis study and using 8 different databases for a thorough search. In addition to this primary objective, the study aims to identify any gaps in the existing literature, such as variations in study design and methodological limitations that may affect the validity of the results. By highlighting these gaps, the study can provide insights into future research directions and suggest ways to address these limitations effectively.

## Materials and Methods

### Eligibility Criteria

We designed this study based on the methodology of the Preferred Reporting Items for Systematic Reviews and Meta-Analyses (PRISMA) guideline (13). The registration number in the International prospective register of systematic reviews (PROSPERO) is CRD42021277901 (Available: https://www.crd.york.ac.uk/prospero/display_record.php?RecordID=277901)

### Inclusion/exclusion criteria

Studies meeting the following criteria were included: 1) human case-control, cohort, and cross-sectional studies reporting on the relationship between thyroid disorders and CD, 2) Reporting of at least one variable showing relative risk effect sizes including risk ratio (RR), odds ratio (OR), and hazard ratio (HR) in both case and control groups. The diagnosis of thyroid disorders in included studies should have been confirmed by serology tests and clinical signs and symptoms. Furthermore, the confirmation of CD should have been based on endoscopy findings, biopsy results, serology tests, and/or HLA DQ2/DQ8 genotyping. Studies that did not include any of this information (including animal studies) were excluded from further analysis.

### Categories of thyroid disorders

Based on a textbook (14) and data extracted from previous original articles, we used four main categories of thyroid disorders for subgroup analysis including hypothyroidism, hyperthyroidism, ATD, and thyroiditis. ATD also had three subcategories including Graves’ disease, Hashimoto’s thyroiditis, and autoimmune hypothyroidism. Some original articles had an emphasis on coexistent hypothyroidism and autoimmunity, but other studies just reported the term ATD; therefore, the association of CD and autoimmune hypothyroidism was also evaluated in the articles that clarified it separately as a subcategory of ATD patients.

Some articles also evaluated and reported the positivity of TPO antibodies and TG antibodies. We extracted and included them in both total and separate analyses.

### Information Sources and Search Strategy

On 28 February 2022, we conducted our systematic search in the following databases to identify relevant articles: PubMed, Web of Science, Scopus, Ovid, Embase, Cochrane, ProQuest, and Wiley. Thorough a three-step process our search keywords and search syntax were designed. First, we extracted the concepts needed for the search following PICO analysis in proportion to the research topic. To accomplish maximum comprehensiveness to recover concepts, we extracted and inserted synonyms, abbreviations, related terms, UK / US spellings singular / plural forms of words, and thesaurus terms in our search syntax. We used the thesaurus MeSH and Emtree for completing keywords. Furthermore, thematic searches on databases were performed. Finally, preparatory search was conducted and related keywords of main articles in this subject were analyzed. We also enriched and completed the vocabulary. Consequently, the baseline syntax was Thyroid AND (Celiac OR coeliac OR Gluten Enteropath* OR Sprue). Complete search strategies on each database are explained in Supplementary 1. Third, two authors hand-scanned the manual reference lists of all studies and relevant systematic reviews to avoid any missing. We also tracked citation for all the included articles.

**Supplementary 1.**
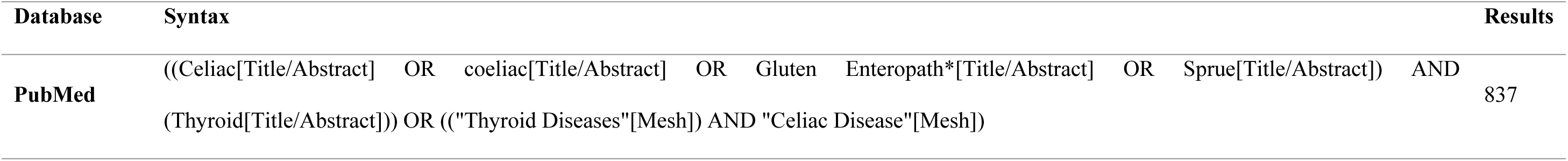
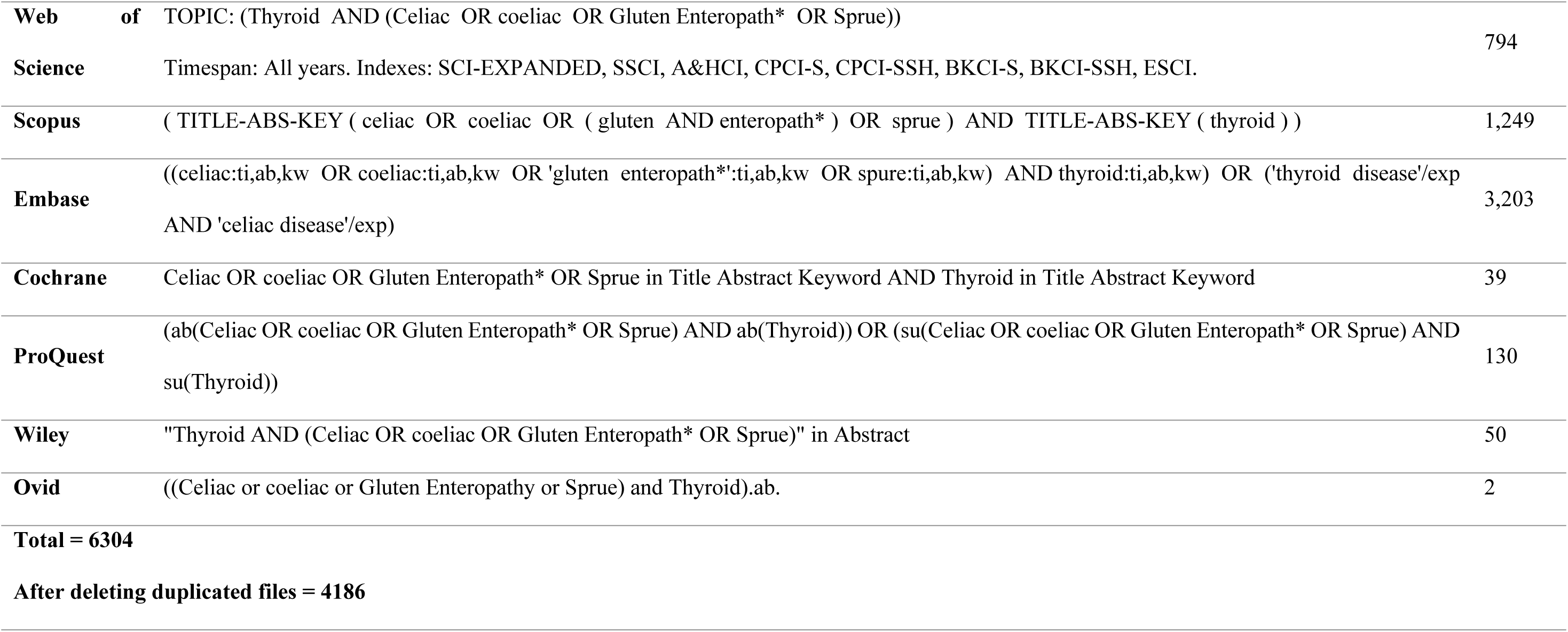

### Quality and risk of bias assessment

To eliminate duplicates and sift through the studies, all articles gathered from the databases were imported into Endnote software (Version X9; Thompson Reuters Corporation, Toronto, ON, Canada). Initially, screening was carried out by assessing the titles and abstracts to determine the eligible articles. Two reviewers (A.Y. and A.K.) independently reviewed the chosen full-texts to classify them into three groups: relevant, irrelevant, and unsure. A third supervisor reviewer (H.S.) resolved any discrepancies. Two independent reviewers (Z.N. and F.H.) evaluated the quality and risk of bias assessment of the eligible articles, using an adapted version of the Newcastle-Ottawa Scale (NOS) form specifically designed for case-control studies. (15). The selected studies were then divided into three groups: poor, fair, and good; according to a score obtained from the selection, comparability, and outcome/exposure categories. This score ranged from zero to eight. In case of any disagreements between the two reviewers, a third reviewer (S.B.) was brought in to resolve the issue.

### Data extraction

Four authors (Z.N, F.H, A.K, and A.Y) extracted the data from each paper based on the list of required variables. We divided the data into three categories: general information, risk of bias assessment, and study setting. The general information category included the first author’s names, year of publication, country, and type of study. The risk of bias assessment category focused on assessing the risk of bias in each study. The study setting category included study duration, sample size, total population recruited in each study, age group and age range, sex, the definition of CD, source of the data, diagnostic methods, each thyroid disorder, P-value, and the ethical approval. To fix missing items or providing additional necessary data, we sent an e-mail to the corresponding authors of those articles, if possible.

### Data synthesis and analysis

We used STATA software, version 12, to calculate pooled OR and its corresponding 95% CIs for each study related to the chance of developing thyroid disease in patients with CD. The combined OR was presented in a forest plot. Heterogeneity among studies was measured using the Q Cochrane test and I^2^ index. A value less than 50% was considered moderate heterogeneity, while a value above 50% was considered severe heterogeneity. According to the existence of heterogeneity in the methodology of the studies, at moderate level of heterogeneity, fixed-effect model was used and at severe levels of heterogeneity, random-effect model was used to combine data in meta-analysis. For finding the cause(s) of heterogeneity, and to investigate the effect of the type of studies and their quality on the research results subgroup and sensitivity analysis was used. Subgroup analysis evaluated factors including the thyroid outcome, type of study, and the risk of bias assessment. In order to evaluate for the presence of any potential publication bias, Begg’s and Egger’s tests were performed to assess asymmetry in the funnel plot. A *P*-value of less than 0.05 was deemed to be significant. To further investigate the impact of individual studies on the overall pooled OR, a sensitivity analysis known as the one-out remove method was utilized. Because two different populations in two different time periods were reported in the Grode et al., 2018 study (3), the data of this study was included separately in the analysis, as data I and II.

## Results

### Search results

In our primary search, 6304 papers were found in the mentioned databases. After removing duplicates, 4186 citations remained for the title and abstract screening. Based on the inclusion and exclusion criteria, 4114 papers were excluded and 72 full-text articles were used for screening. Out of the available full-text studies, 31 were deemed eligible and included in both the systematic review and meta-analysis. The reasons for excluding other full-text studies are listed in Figure 1.

**Figure 1.**
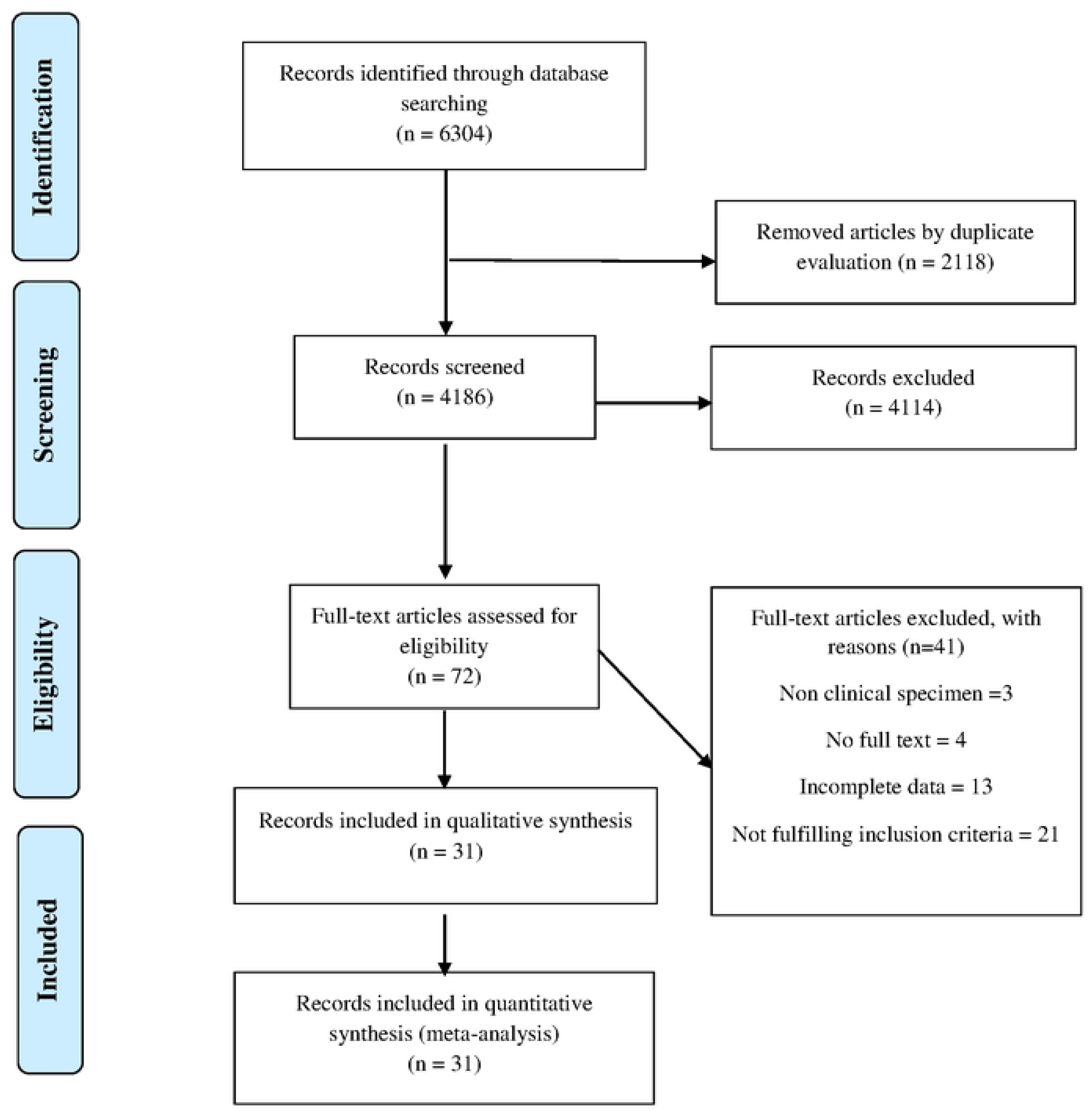
The process of study selection.

### Characteristics of the included articles

Table 1 shows the detailed description of key characteristics for the 31 included studies. There were 3310256 participants in total, of these, 101253 were in the thyroid group and 3209003 were in the non-thyroid group. Among the studies, Nafeesa et al. had the highest study population with 2426224 participants, including 6506 cases and 2419718 controls (16). All studies were published between 1998 and 2022; furthermore, the data of studies were from the Europe and Asia continents, as well as the USA. In 47.06% of the studies, the thyroid was defined as autoimmune. As a results, the NOS assessment showed that only 67.74% of the studies had good quality.

**Table 1.**
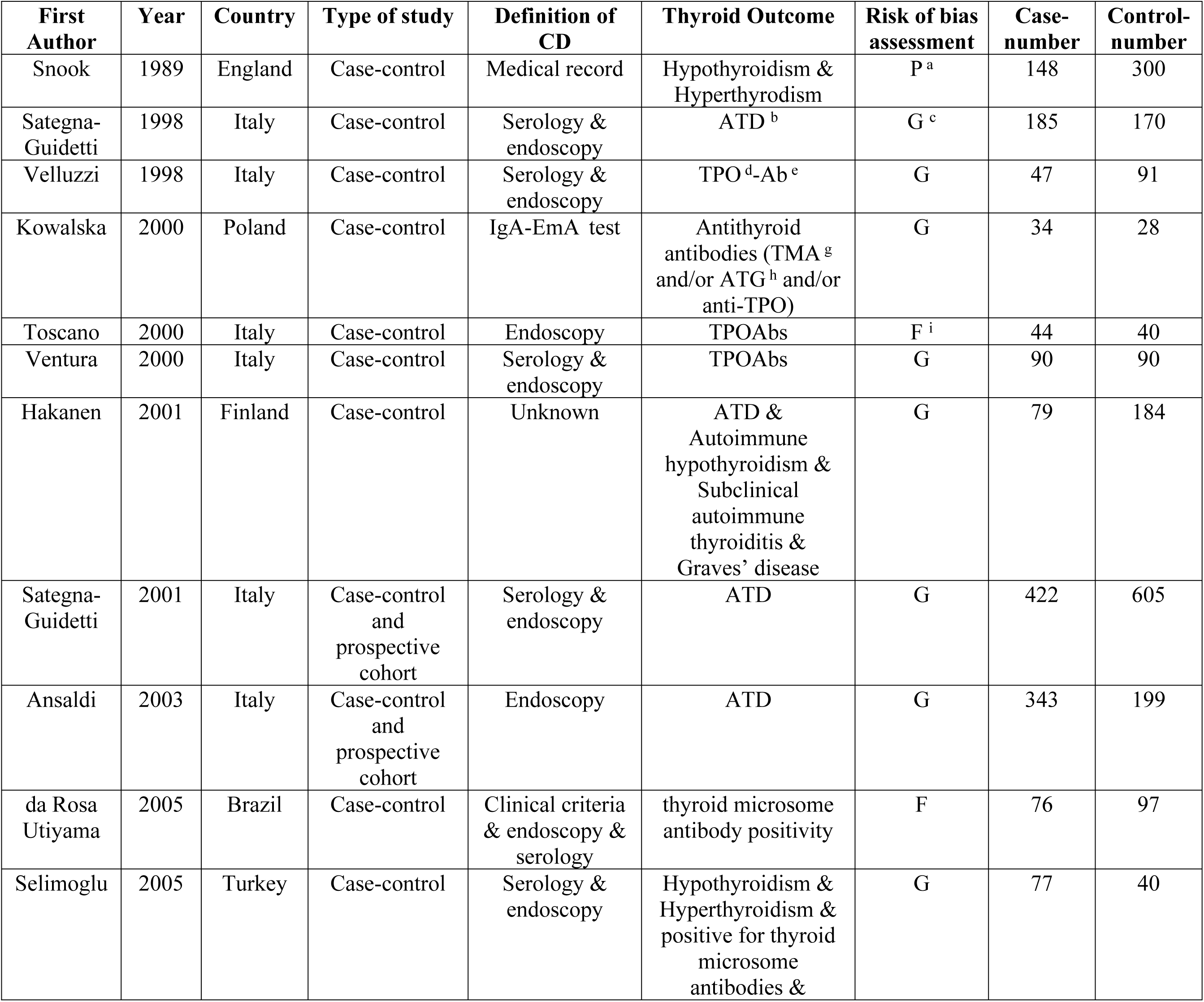
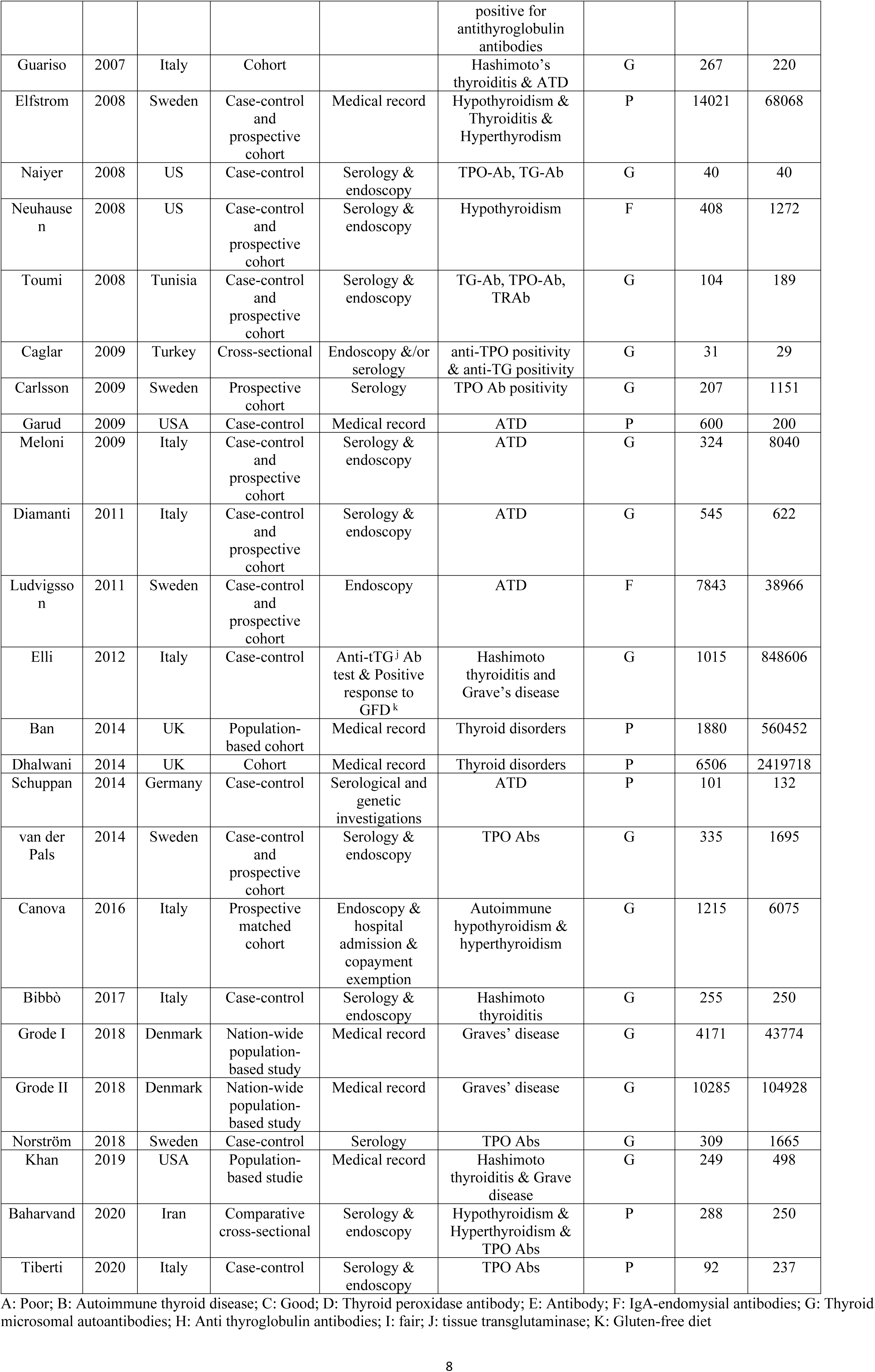
Main characteristics of the included studies in the meta-analysis on the thyroid disorders infection and celiac disease (CD)

The meta-analysis demonstrated that there is a significant relationship between CD and thyroid disorders (pooled OR: 3.06; 95% CI: 2.51 - 3.72; P ≤ 0.001). Forest plot of the frequency of thyroid disorders in CD patients compared with the control group is shown in Figure 2.

**Figure 2.**
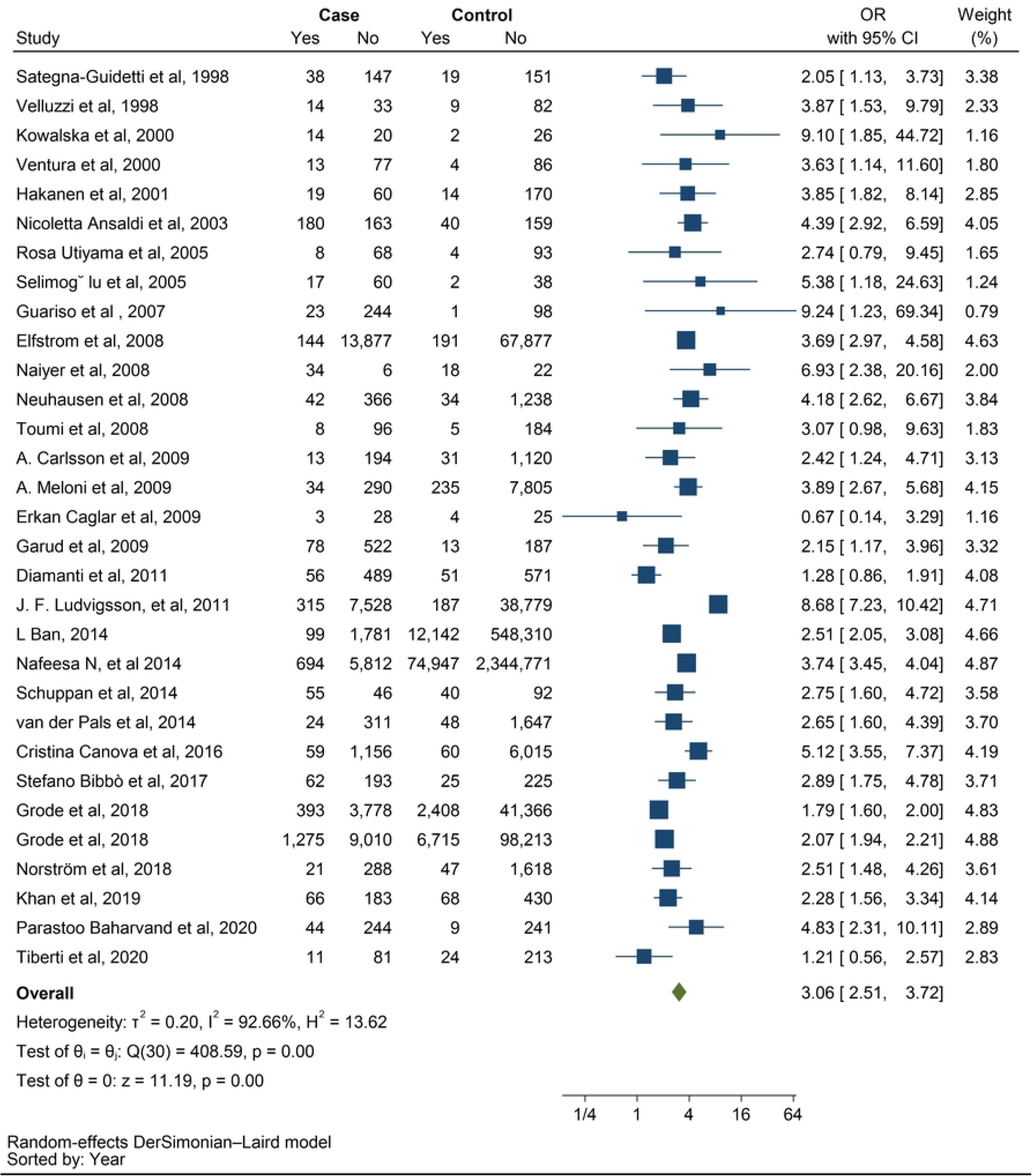
Forest Plot illustrating the strong association between Celiac Disease (CD) and thyroid disorders (p < 0.001, pooled OR: 3.06; 95% CI: 2.51 - 3.72).

### Subgroup analysis

To assess the heterogeneity of the studies, we conducted multiple subgroup analyses based on three factors: types of thyroid disorders, types of studies, and quality of studies. The amount of heterogeneity did not change significantly within the subgroup analyses, therefore they cannot be a source of the heterogeneity. As shown in figure 3, In the subgroup analysis by study type, cross-sectional studies had a lower pooled OR than case-control and cohort studies (OR: 2.04 vs OR: 3.09 and OR: 3.48, respectively).

**Figure 3.**
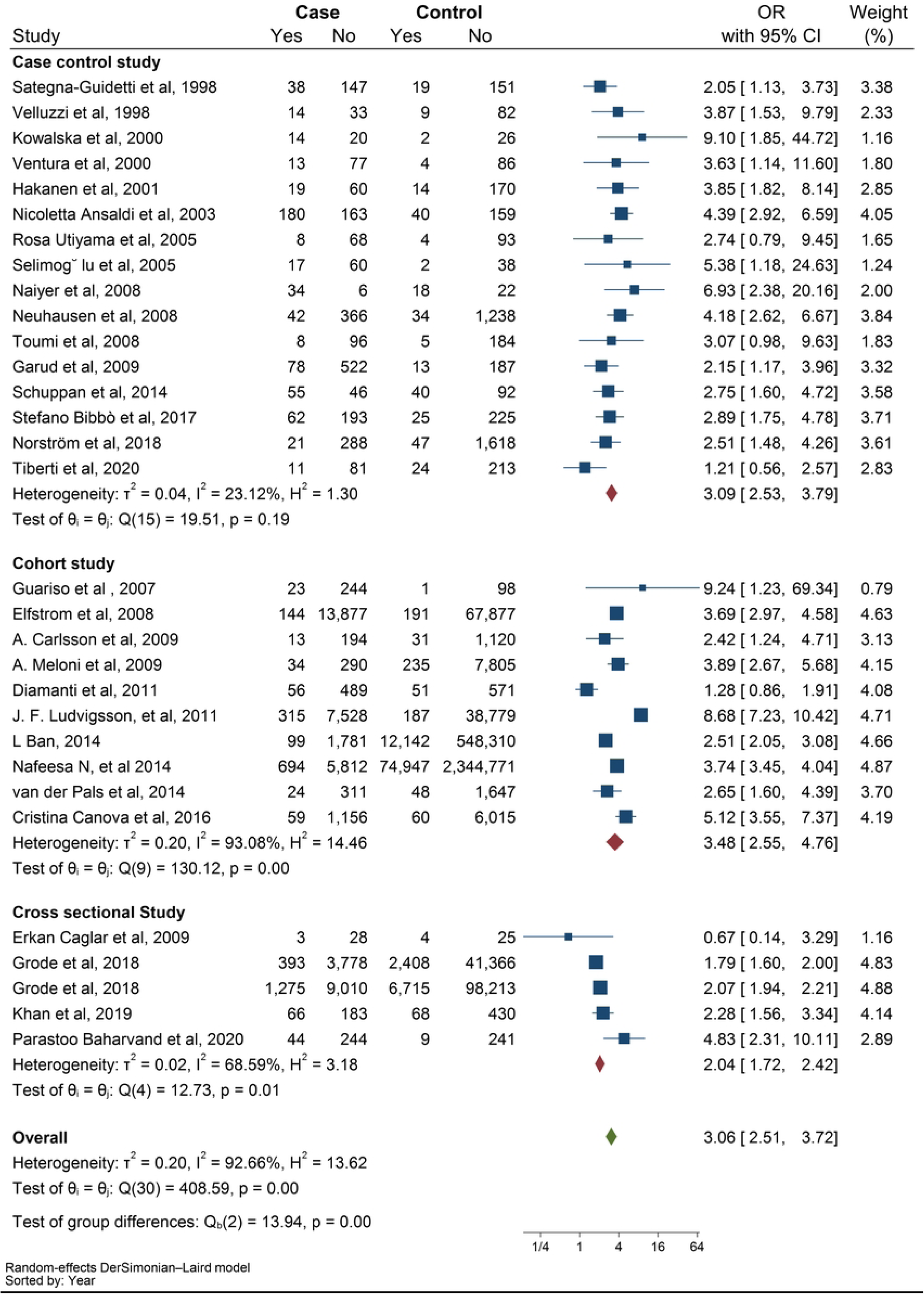
Demonstration of subgroup analyses based on type of study.

**Figure 4.**
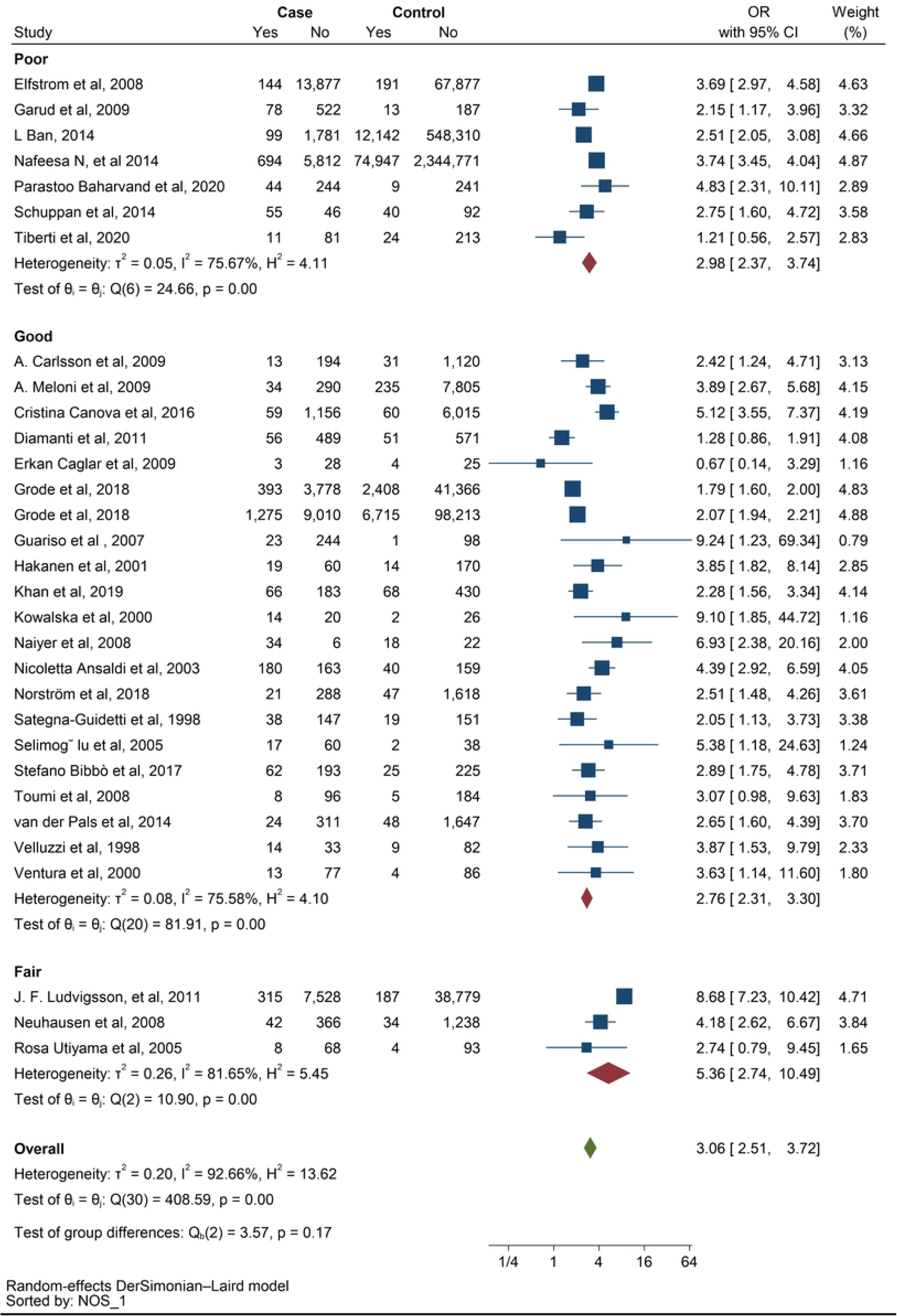
Subgroup analysis based on Newcastle-Ottawa Scale (NOS) assessment.

Based on the NOS assessment, most studies had good scores. The pooled OR for fair studies were higher than poor and good studies (OR: 5.36 vs OR 2.98 and OR: 2.76 respectively).

Furthermore, the analysis showed a significant relationship between CD and ATD (OR: 2.96, CI: 2.32 – 3.78, P ≤ 0.001). (Figure 5).

**Figure 5.**
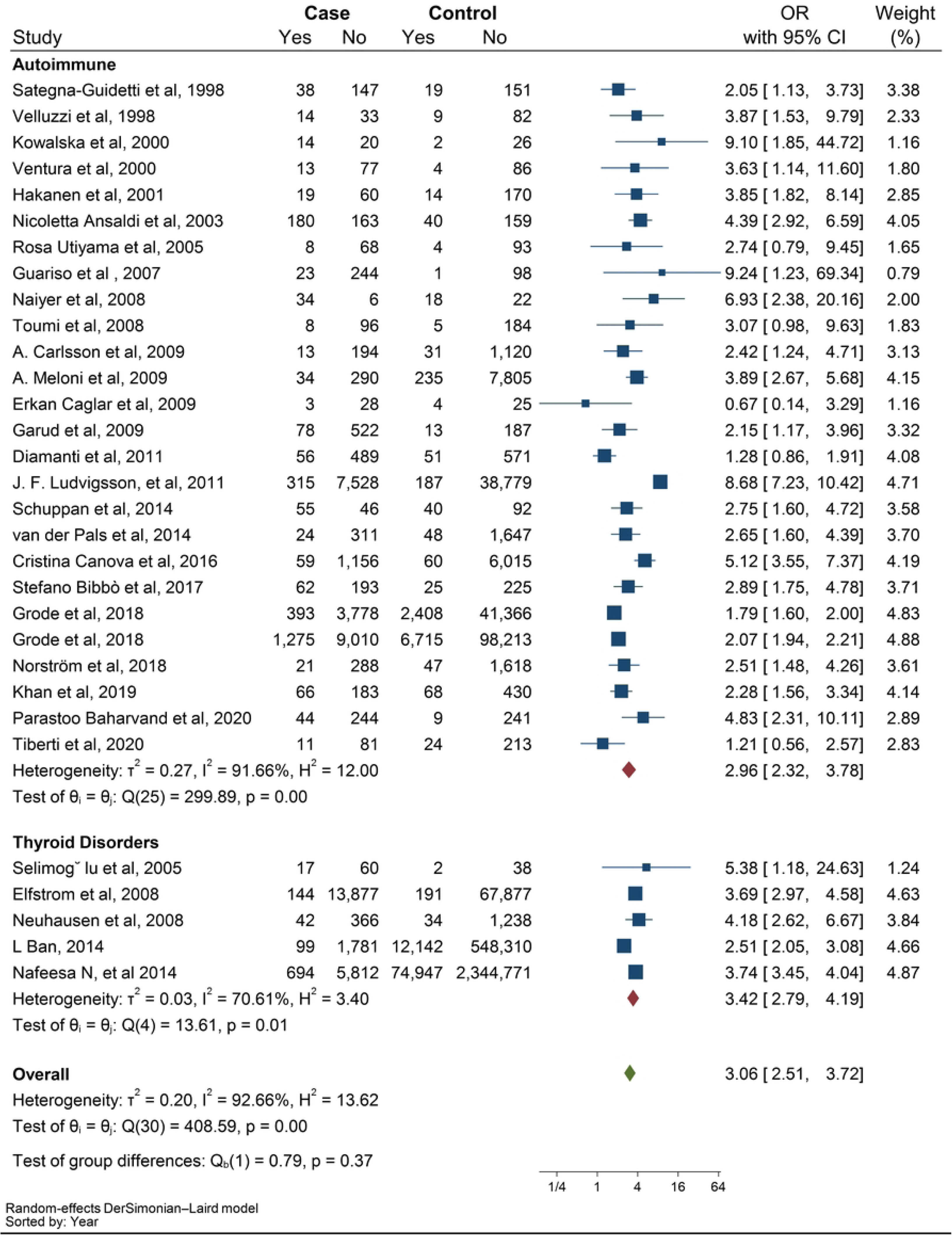
Subgroup analysis based on autoimmune thyroid disease (ATD) and other thyroid disorders.

Random effects meta-analysis was conduct for interaction in planned subgroups and adjust common effects (Figure 6).

**Figure 6.**
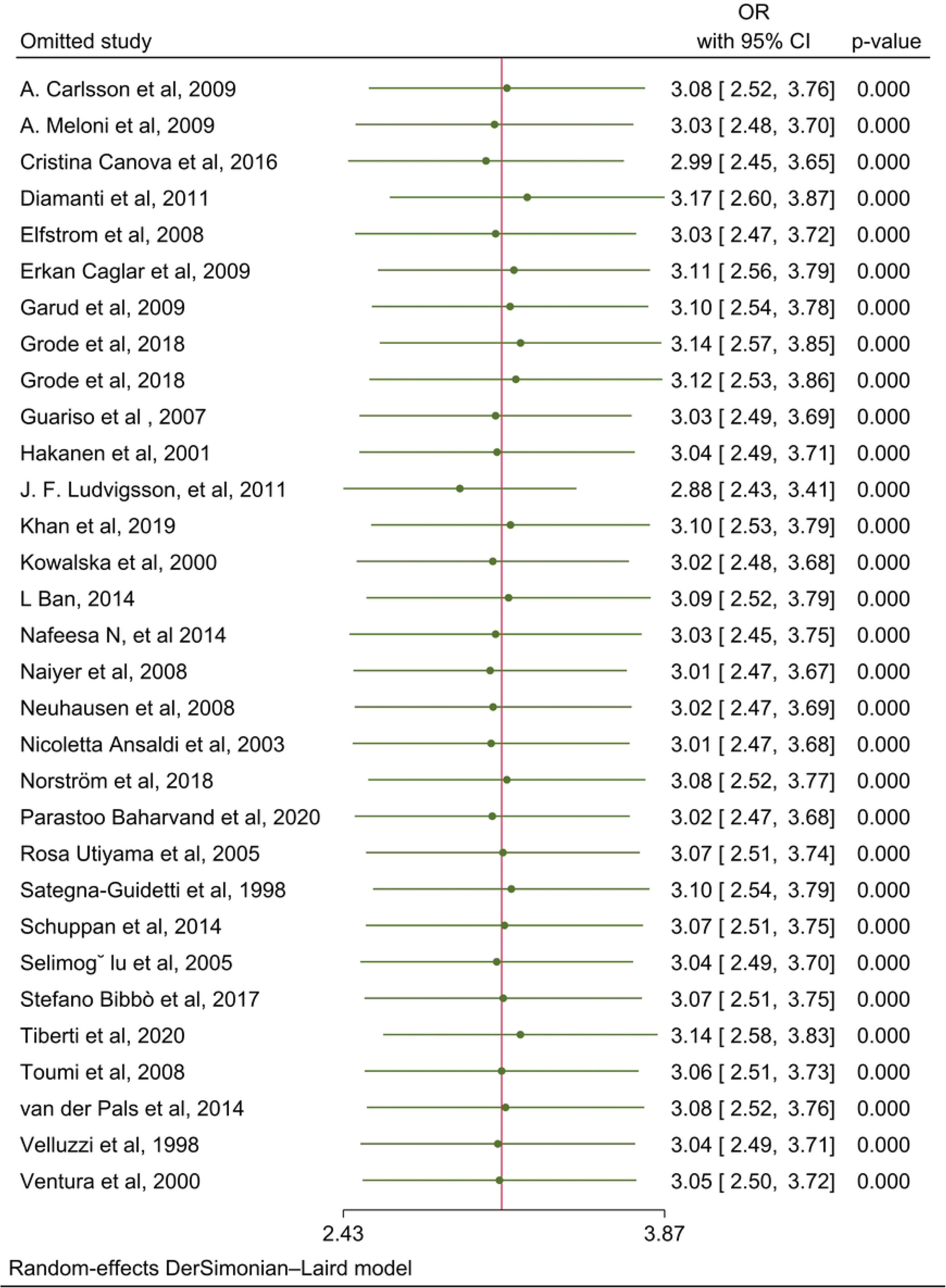
Random effects of all 31 articles.

### Publication bias

The results of Begg’s and Egger’s tests indicate that the funnel plot does not exhibit significant asymmetry, suggesting the absence of publication bias in the data. This inference is supported by the p-values obtained from Begg’s test (0.75) and Egger’s test (0.61) (Figure 7).

**Figure 7.**
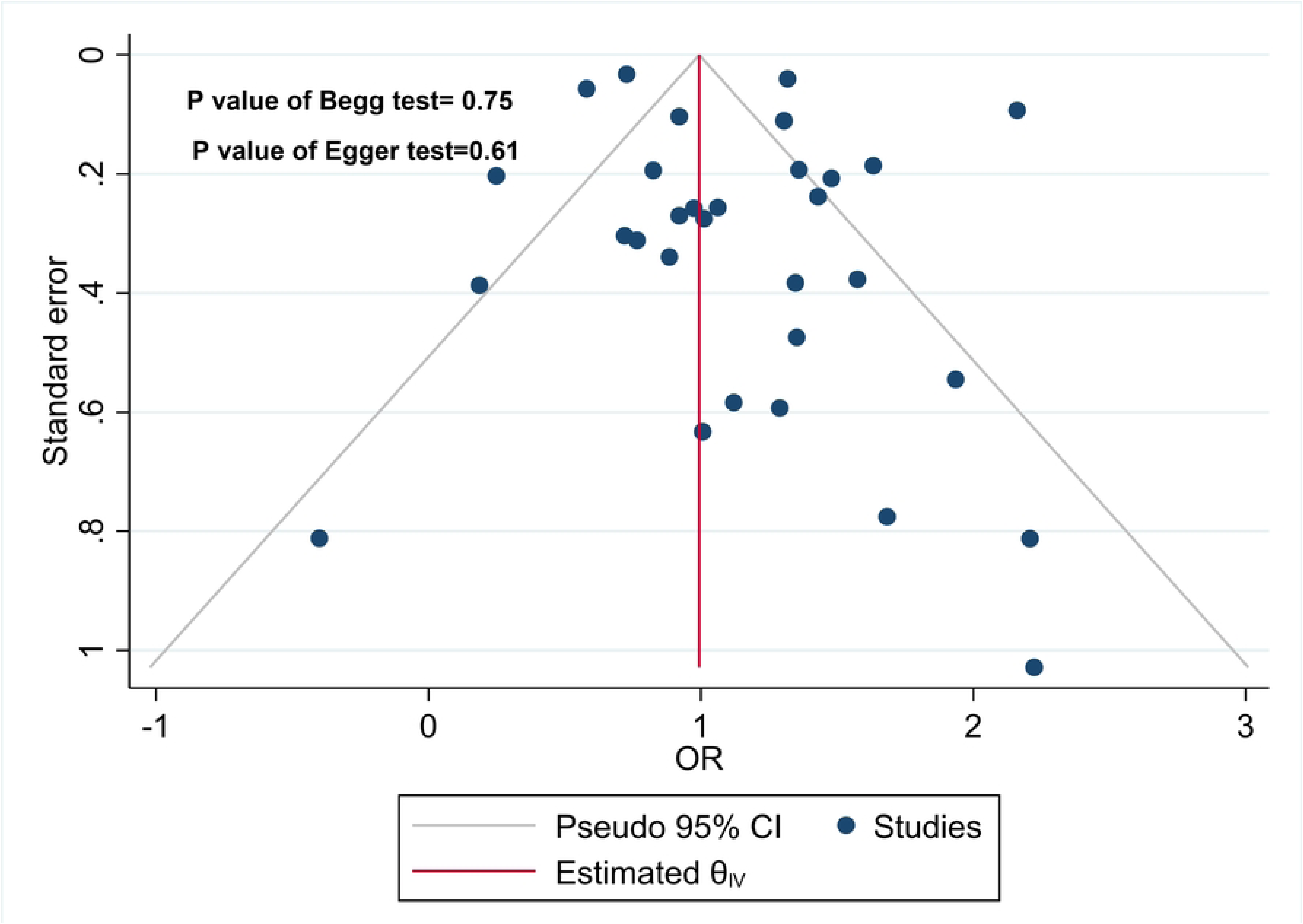
Begg’s test (p = 0.75) and Egger’s test (p = 0.61) results confirm the absence of significant publication bias.

## Discussion

To evaluate the risk of thyroid disorders in patients with CD, several case-control and cohort studies have been conducted or are currently underway. However, these studies have varying methodologies and sample sizes, and their results are inconclusive. To address this issue, a systematic review and meta-analysis was conducted to combine all relevant studies and establish strong evidence of any association between CD and thyroid disorders. After analyzing the findings of 31 studies, we have demonstrated that individuals with CD are at a significantly higher risk of developing thyroid disorders compared to those in the control population. In fact, the risk is found to be 3.06 times greater for CD patients.

The pathophysiologic reason why thyroid disorder is more common with CD are still under discussion and are not well known. Regarding that, there are several suggested mechanisms. One of them is related to genetic factors. Studies have shown that human leukocyte antigen (HLA) is overexpressed in celiac disease. Specifically, HLA-DQ2 and DQ8 have been found to be associated with ATDs, particularly Hashimoto’s thyroiditis. A meta-analysis conducted on this topic revealed that Hashimoto’s thyroiditis was more than three times as prevalent in CD patients compared to those without CD. The gene that encodes cytotoxic T lymphocytes is related to antigen 4 (CTLA-4) and ATD. CTLA-4 is located in the HLA region and is also associated with CD, therefore can have a strong connection with thyroid autoimmune diseases (17, 18, 19).

In CD, patients suffer from malabsorption of nutrients such as iron, selenium, and vitamin D, due to chronic inflammation and villous atrophy. Iron deficiency worsens thyroid dysfunction due to the decreased activity of the heme-dependent TPO, which relates to ATD (e.g., Hashimoto’s thyroiditis and Graves’ disease) (20). Another involved mechanism is the deficiency of selenium. This deficiency can modulate the expression of selenium proteins, resulting in the inflammation and mucosal damage. Selenium deficiency can negatively impact thyroid function, since selenium proteins act as antioxidants and regulate redox status and thyroid hormone metabolism in thyrocytes. The selenium proteins are also involved in cell growth and apoptosis (21). The mechanism related to vitamin D deficiency and thyroid disorder can be explained as follows: due to vitamin D deficiency, the suppression of T cells is disrupted, resulting in an increase in the release of inflammatory cytokines. This leads to the destruction of thyroid tissue and ATD (22).

Additionally, tTG-2 IgA antibodies have also been proven to react with thyroid tissue, which may contribute to thyroid disease in CD. As Naiyer et Al. observed a positive relation between tTG-2 IgA and anti-TPO antibody titers, which indicated the relationship between CD and the presence of organ-specific antibodies against thyroid tissue (23).

A meta-analysis study conducted in 2016 found that thyroid disorders are three times more prevalent among CD patients compared to those in the control group (OR 3.08, 95% CI 2.67-3.56; P<0.001). In their subgroup analysis for hyperthyroidism by three original articles, they demonstrated that hyperthyroidism has no significant association with CD. However, when a subgroup analysis was conducted using five articles in the present study, it revealed that CD patients have a two-fold risk of developing hyperthyroidism, similar to the impact of the other types of thyroid disorders. On the other hand, our subgroup analysis for hypothyroidism was consistent with the last meta-analysis. The evaluation revealed that the occurrence of hypothyroidism in CD patients was significantly higher when compared to individuals in the control groups (OR 3.38, 95% CI 2.73–4.20, P<0.001) (12).

After the final search, we reached 18 articles more than the last systematic article. According to new articles and additional data, the categories of thyroid disorders were increased in our study and we analyze them in separate subgroups to reach their specific OR. We also, analyzed the studies according to groups of the quality of the studies based on NOS assessment and types of studies; to determine the possible discrepancies. Fortunately, no discrepancies were detected. Finally, all subgroup analyses demonstrated a strong effect of CD on the occurrence of different thyroid disorders.

However, there are some limitations in this study. Some original articles specified that the patients had euthyroid autoimmune disease or autoimmune hypothyroidism, but other articles did not mention that. We suggest for future original articles to distinguish the type of autoimmunity. The other limitation is that low number of studies from Asia, and lack of researches from Africa, South America, and Oceania. So that, the results more emphasize the population in America and Europe. Evaluating other ethnicities is helpful for further studies.

### Conclusions

Our systematic review and meta-analysis of 34 original articles included 102545 CD patients show that they are at three to four-fold risk of thyroid disorders, especially autoimmune hypothyroidism. We suggest that a design of a cohort study with large sample size of CD patients for a long-time follow-up to evaluate any possible thyroid disorders. Furthermore, based on the findings, it is essential to conduct routine screening for thyroid disorders in individuals with CD.

## Data Availability

All relevant data are within the manuscript and its Supporting Information files.

N

## Abbreviations

CD: Celiac disease
OR: Odds ratio
CI: Confidence interval
ATD: Autoimmune thyroid disease
TPO: Thyroid peroxidase
TG: Thyroglobulin
TSH: Thyroid-stimulating hormone
PRISMA: Preferred Reporting Items for Systematic Reviews and Meta-Analyses
RR: Risk ratio
HR: Hazard ratio
NOS: Newcastle-Ottawa Scale
CTLA-4: Cytotoxic T lymphocytes is related to antigen 4

## Acknowledgements

Not applicable.

## Authors’ contributions

Z.N: supervise the project; data extraction; drafting of the article. F.H: helped supervise the project; data extraction; drafting of the article. S.B: conception and design; revision of the article. H.Sh: supervise the project; review screening the articles. S.Gh: data analysis; revision of the article. A.Y: screening the articles; data extraction. M.M: search strategy and writing syntax; revision of the article. A.K: screening the articles; data extraction. All authors discussed the results and contributed to the final manuscript.

## Availability of data and materials

All data generated or analyzed during this study are available.

## Declarations

### Ethics approval and consent to participate

Not applicable.

### Consent for publication

Not applicable.

